# Functional network dynamics in photosensitive epilepsy depend on stimulation frequency and photosensitivity type

**DOI:** 10.1101/2024.12.21.24319242

**Authors:** Lili Timar, Sina Deplazes, Julia Bothmann, Roland Renzel, Debora Ledergerber, Tena Dubcek, Lukas Imbach

## Abstract

**Objective:** To describe the underlying dynamical mechanisms of photosensitive epilepsy (PSE) and different photosensitivity types.

**Methods:** This retrospective study included healthy controls (HC), non-PSE patients with epilepsy (PWE), and PSE patients. All participants were stimulated with flickering white light (1-60Hz) under 10-20 EEG monitoring.

**Results:** We observed significantly higher occipital Photoparoxysmal response (PPR) power in PSE patients compared to HC for stimulation frequencies 10Hz-20Hz. This activity was highest in Type 4, which shows a type-specific variation of the PPR. However, the excitability of the visual cortex, investigated by the occipital transient (P100) and steady-state visual evoked potentials (ssVEP), remained unchanged in most PSE types compared to HC and to PWE. The ssVEP power only increased significantly in Type 4 compared to HC. Instead, PSE patients showed apparent differences in functional connectivity in the PPR band (3-4Hz) with hypoconnected centro-parietal regions and hyperconnected anterior and anterio-posterior areas compared to HC and PWE for stimulation frequencies 10Hz-20Hz.

**Conclusion:** PSE is a network effect modulated by hyperconnected anterior and anterio-posterior regions, accompanied by a hyperexcitable visual cortex.

**Significance:** We provide novel evidence that altered network dynamics in PSE patients are likely a prerequisite for the propagation of the PPR and type-specific responses.

## 1. Introduction

Photosensitive epilepsy (PSE) affects 5% of epilepsy patients worldwide, with a high prevalence in females, children, and adolescents (Fisher et al., 2022; Martins da Silva & Leal, 2017). It is diagnosed via the Intermittent Photic Stimulation procedure (IPS) under concurrent Electroencephalography (EEG) monitoring (Kasteleijn-Nolst Trenité et al., 2012). The IPS delivers white flashes of light that evoke transient visually evoked potentials (tVEP) for lower and steady-state visual evoked potentials (ssVEP) for higher stimulation frequencies. Both the tVEP and the ssVEP are normal physiological reactions, time-locked to the stimulus and linked to the triggering frequency (Herrmann, 2001). However, stimulation frequencies specific to PSE patients, mainly between 15 and 20 Hz, can induce abnormal EEG activity, characterized by 3-4 Hz spikes or spike-slow wave complexes, called the photoparoxysmal response (PPR) (Fisher et al., 2005; Harding & Harding, 2010; Kasteleijn-Nolst Trenité et al., 2012; Taoufiqi et al., 2016). Based on the propagation of the PPR, PSE can be further categorized into four different types ranging from least to most severe (Type 1-4) (Waltz et al., 1992). Photosensitivity occurs mainly in genetic (idiopathic) generalized epilepsies (GGE) but also in focal idiopathic photosensitive occipital epilepsy (IPOE) and has a high prevalence in certain generalized epilepsy syndromes such as juvenile myoclonic epilepsy (JME) or juvenile absence epilepsy (JAE) (Guerrini & Genton, 2004; Martins da Silva & Leal, 2017; Taylo et al., 2013). Both focal (occipital) and generalized (whole brain) network effects have been hypothesized and investigated as the driving mechanism of PSE (Brinciotti et al., 2020a; Moeller et al., 2013; Strigaro et al., 2012; Varotto et al., 2012).

Previous studies in PSE have revealed increased amplitude of the occipital tVEP, the so-called P100, in response to single- and paired flashes compared to healthy controls (HC) (Brinciotti et al., 2020; Strigaro et al., 2012). It was suggested that augmentation of the P100 amplitude indicates a focal hypersynchronization of the occipital cortex, potentially leading to hyperexcitability and seizures (Wilkins et al., 2004). Further indication of a hyperexcited visual cortex associated with PPR was shown in a study using transcranial magnetic stimulation that found lower phosphene thresholds in individuals with PPR compared to those without (Siniatchkin et al., 2007a). Recent papers have also identified abnormal activity of specific EEG frequency bands in the visual cortex of PSE patients in the form of increased alpha and gamma oscillations, highlighting its altered excitatory and inhibitory processes (Hermes et al., 2017; Vaudano et al., 2017).

While the focal hyperexcitability of the visual cortex has been associated with PSE, the extent to which it is causally involved in generating and propagating the PPR is still debatable. Moeller et al. (2009a) found that during a PPR, blood oxygen level-dependent (BOLD) responses increased in the occipital cortex of PSE patients, however, similar increases were observed in the parietal and frontal regions. In a later study, Moeller et al. (2013) proposed that PPR is a cortical phenomenon and modeled the general network of its propagation to involve occipital, parietal, and frontal areas. Similarly, a connectivity study in the EEG-gamma band during photic stimulation pointed towards hyperconnectivity as the facilitator of the PPR. They showed higher anterior functional connectivity, suggesting that the PPR might depend on hyperconnected frontal regions (Varotto et al., 2012). These findings indicate that the hyperexcitability of the visual cortex is a concomitant rather than the primary driver of the PPR and that network-based effects are responsible for PSE.

Here, we aimed to solidify the role of the network in PSE and to provide a more detailed understanding of its underlying dynamics. We retrospectively analyzed scalp EEG recordings under IPS stimulation of PSE patients (Type 2, Type 3, and Type 4), non-PSE epilepsy patients (PWE), and healthy controls (HC). We assessed the activation of the visual cortex by analyzing the physiological (P100 and ssVEP) and pathological (PPR) response patterns in reaction to IPS. Additionally, the propagation characteristics of the PPR were investigated by evaluating the functional connectivity (FC) in the low-frequency band (3-4 Hz), where pathological activity occurs but so far remained unexplored.

## 2. Methods & Materials

Our aims for the present study were to (i) characterize type- and stimulation frequency-specific PPR activity in PSE groups, (ii) directly assess and compare the activity of the visual cortex in response to IPS in PSE, non-PSE, and HC groups, (iii) map dynamic FC patterns of PSE patients in response to high-risk stimulation frequencies in the PPR band.

### 2.1 Study populations

We retrospectively reviewed a large cohort of epilepsy patients and healthy participants (HC) who underwent a routine EEG recording at the Swiss Epilepsy Center. All participants were stimulated with IPS under EEG monitoring along standardized protocols according to the International Federation of Clinical Neurophysiology (IFCN) guidelines (Peltola et al., 2022). Recordings were carried out as routine EEG patient follow-ups and as occupational fitness assessments for the HC. Two blinded independent medical professionals evaluated the encrypted EEG recordings and determined the PSE Type (2, 3, or 4) or lack thereof (PWE). A third medical professional provided the final classification in case of differing classification. The patients’ epilepsy syndromes were categorized based on the International League of Epilepsy (ILAE) criteria (Berg et al., 2010; Hirsch et al., 2022), into genetic generalized (GGE), focal (FE: structural, non-structural or unknown), combined (CM: generalized and focal) or epilepsy of unknown origin (UC).

We selected 45 PSE patients who fulfilled the criteria of Type 2 (n = 11; mean age 29.5 ± 16.5, 10 females), Type 3 (n = 14; mean age 22.8 ± 12.9, 12 females), or Type 4 (n = 20; mean age 21.1 ± 8.6, 19 females). Photosensitivity was determined by the observation of the PPR under IPS stimulation. Occipital spikes and slow waves with parietal propagation were categorized as Type 2, parieto-occipital spikes and slow waves with spread to the frontal regions as Type 3, and generalized spike and wave complexes as Type 4 (Waltz et al., 1992). Type 1 patients were excluded due to frequent ambiguity with PSE Type 2.

We also recruited 19 non-PSE patients with epilepsy (PWE, mean age 22.2 ± 8.7, 15 females) and 20 healthy individuals (HC, 21 ± 2.7, 3 females) as control groups. The PWE group consisted of epilepsy patients who, under IPS stimulation, exhibited no PPR nor sensitivity upon eye closure, indicating no photosensitivity. Finally, individuals of the HC group exhibited no PPR under IPS stimulation nor sensitivity upon eye closure and had no current or history of epilepsy. Additionally, the participants in this group took no CNS active medication nor had cognitive deficits. For the categorization of the patients’ (PSE, PWE) epilepsy syndromes and ASM intake, refer to Table 1.

**Table 1.:**
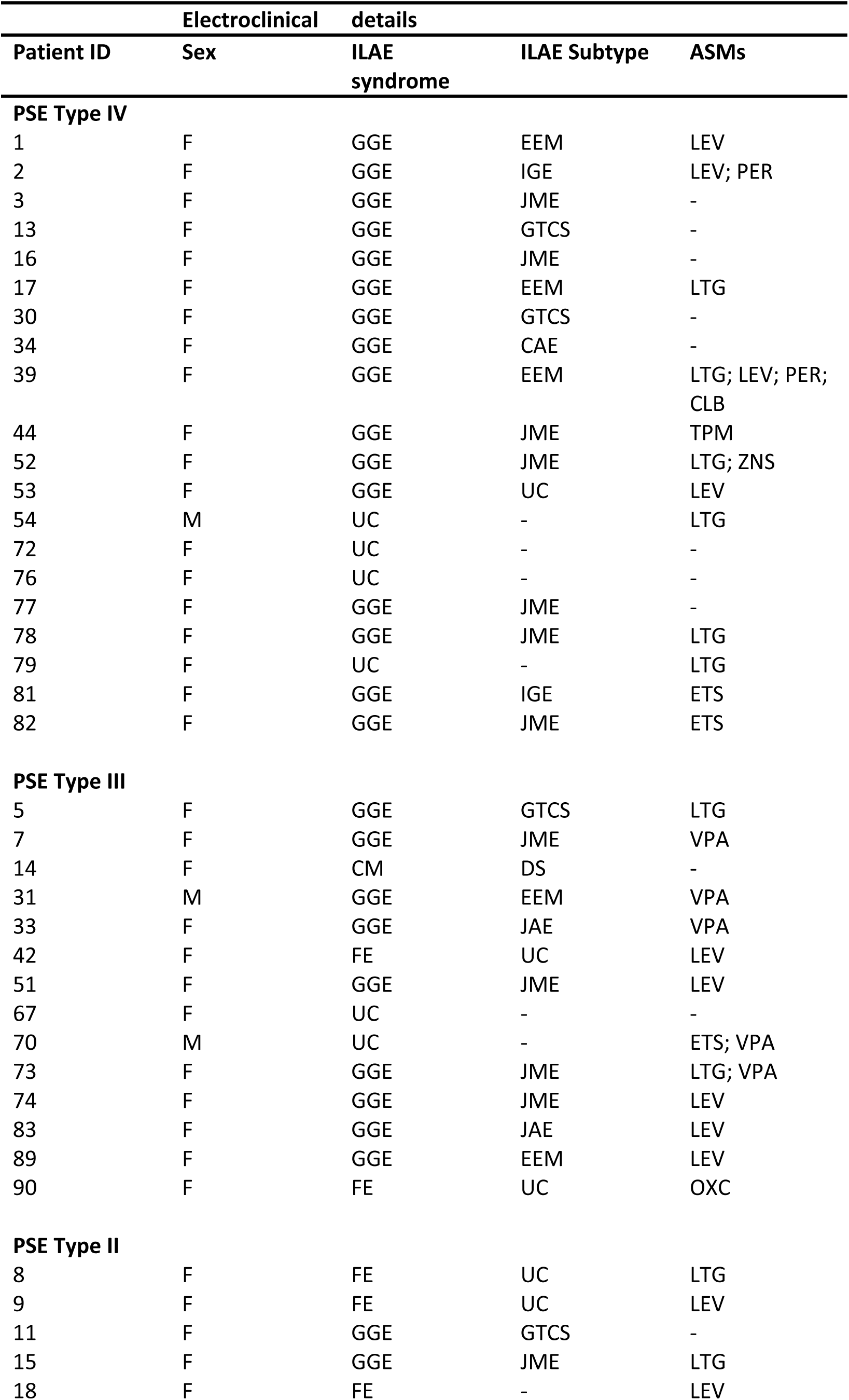
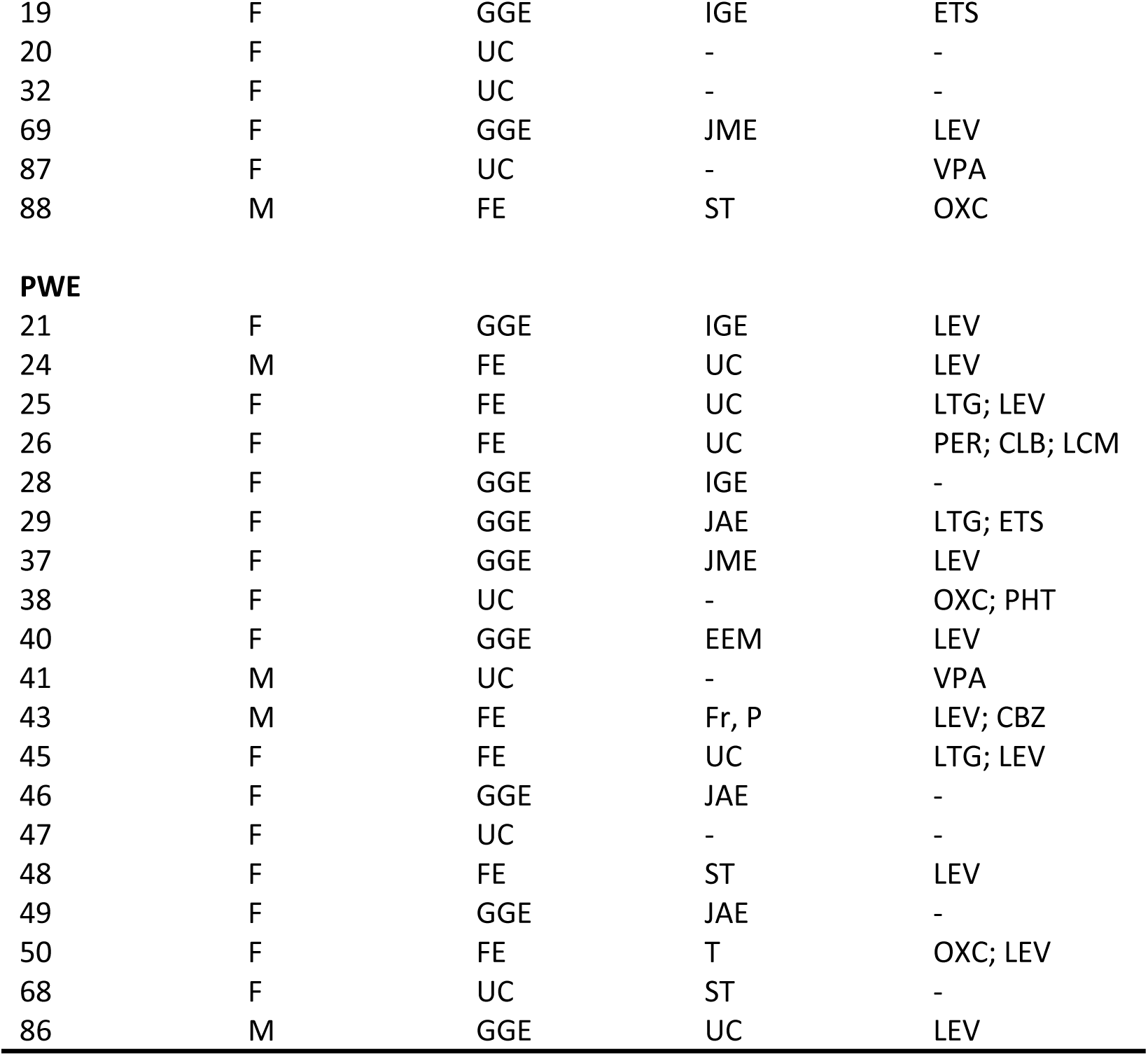
Electroclinical details and demographics of the patient groups. ASM = antiseizure medication; CAE = childhood absence epilepsy; CBZ = carbamazepine; CLB = clobazam; CM = combined; DS = doose syndrome; ETS = etosuccimide; EEM = eyelid myoclonia; F = female; Fr = frontal; FE = focal epilepsy; GGE= genetic generalized epilepsies; GTCS = generalized tonic clonic seizures; IGE = idiopathic generalized epilepsy; ILAE = International League Against Epilepsy JAE = juvenile absence epilepsy; JME = juvenile myoclonic epilepsy; LCM = lacosamide; LEV = levetiracetam; LTG = lamotrigine; M = male; OXC = oxcarbamazepine; P = parietal; PER = Perampanel; PHT = phenytoin; PWE = Patients with epilepsy; ST= structural; T = temporal; TPM = topiramate; UC = unclear; VPA = valproate; ZNS = zonisamide

All subjects’ consent was obtained according to the Declaration of Helsinki and was approved by the local ethical committee (Kantonale Ethikkommission, KEK Zurich, KEK number 2024-00401).

### 2.2 EEG recording

EEG was recorded with 23 sintered Ag/AgCl scalp EEG electrodes under the 10-20 system with a low-pass filter of 70 Hz. An additional pericentral electrode was used as reference located close to Cz. Electrode impedance was kept between 2-5 KΩ. For most participants (n=73), the recordings were captured with the NicOne System (NicoletOne™ 44 channel amplifier, Natus, United States) or with the Micromed Ambulatory System (Micromed® Evolution Plus, 40 channels). The sampling frequency was 256 Hz. EEG recordings were conducted in a suitably equipped room at the Swiss Epilepsy Center, Zürich, Switzerland. All participants were lying in a darkened room and completed an approximately 20-minute-long recording with three conditions: resting state (with eyes closed and eyes open), hyperventilation, and IPS (with eyes closed and eyes open). An EEG specialist verbally instructed eye closure and opening, as well as the rate of hyperventilation. During classification, the whole recording was evaluated. However, the analysis in this present study only considered the IPS condition. The IPS stimuli were delivered in four different sections according to IFCN guidelines (Peltola et al., 2022). The participants’ physical state and EEG recording were constantly monitored by the EEG specialists present. IPS was stopped after observing a PPR response or at the participant’s request.

### 2.3 EEG signal preprocessing steps

EEGs recorded with the Nicone System were reviewed with Study Room (NicoletOne LTM version 5.95.0.2) and exported in .edf format, while EEGs recorded with Micromed were reviewed with Brain Quick (System Plus Software version 1.02.0002). For further analysis, 19 channel EEG recordings were loaded with Python (version 3.12) (Python Software Foundation) from .edf or .trc formats, respectively. We applied a high-pass filter of 0.01 Hz to remove baseline wander or drift caused by movement, respiration, or some impedance changes of the electrode. This chosen cutoff frequency also allowed us to observe activity following a stimulation frequency of 1 Hz. Notch filters were used to filter out line noise at 50 Hz. The average of all electrodes served as a reference for all further analyses. Triggers linked to the IPS stimulation carried annotations on the exact stimulation frequencies given throughout the EEG recording. The following stimulation frequencies were consistently recorded for all included individuals: 1Hz, 5Hz, 10 Hz, 15 Hz, 20 Hz, and 25 Hz. Hence, our analyses considered responses to these five conditions.

### 2.4 Visual Evoked Potentials (P100)

We calculated the transient visual evoked potentials (tVEP) for the 1Hz condition by creating 1s epochs, 500 ms before and after every single stimulus, and averaging those for each individual. Automatic rejection of epochs with amplitude greater than +/- 100 μV was applied to reduce artifacts and interferences related to the PPR (Brinciotti et al., 2020). Based on this, two recordings (one Type 4 and one PWE) were excluded from this analysis. The tVEP has two main parts: the main complex, the positive peak (P100), and the after-discharge (Strigaro et al., 2012). In previous studies, the amplitude of the P100 showed significant differences between PSE and non-PSE participants (Brinciotti et al., 2020; Siniatchkin et al., 2007b; Strigaro et al., 2012). Hence, we calculated the peak-to-peak amplitude of the P100 wave and its latency as an additional measure. For group comparisons, we estimated grand averages of the P100; however, for statistical analysis, we considered the individual amplitude and latency values. In nine patients (five Type 4, two Type 3, and one Type 2), 1 Hz stimulation was unavailable (i.e., not recorded). Hence, these individuals were not included at this stage of the analysis.

### 2.5 Power Spectral Density

Power spectral densities (PSD) were calculated on recording segments in response to the remaining stimulation frequencies (5Hz, 10Hz, 15Hz, 20Hz, and 25Hz). We used the fast Fourier transformation (FFT) with a 2s window size, which provided PSD values in a frequency window of 0-60 Hz in bins of 0.5 Hz. We computed PSD at occipital (O1 and O2 averaged), parietal (P3, P4, Pz averaged; in supplementary material), and at frontal electrodes (F3, F4, Fz averaged; in supplementary material) for every individual since these were previously shown to be affected by PPR (Moeller, et al., 2009a).

For each stimulation frequency, spectral power at the fundamental ssVEP band (i.e., the narrow band corresponding to the stimulation frequency) was calculated by the sum of EEG power at three bins respective to the stimulation frequency (4.5-5.5 Hz, 9.5-10.5 Hz, 14.5-15.5 Hz, 19.5-20.5 Hz, 24.5-25.5 Hz) and multiplying that by 0.5 (bin size), resulting in a total power value. The same calculation was done for each stimulation frequency to estimate PPR power, except here, we summed the EEG power in the three bins of the PPR frequency band of 3-4 Hz (3 Hz, 3.5 Hz, and 4 Hz).

There were three patients without a 25 Hz recording (two Type 4, one Type 2) and one Type 2 patient without a 5 Hz recording. Hence, these patients were excluded from this part of the analysis.

### 2.6 Scalp-scalp Functional Connectivity

FC responses to the stimulation frequencies of interest (5Hz-25Hz) were measured by calculating the Phase Locking Value (PLV) with the same segmentation parameters as for the PSD analysis. PLV is suitable for investigating task-induced changes measuring connectivity by estimating the phase-locking consistency of two signals across multiple trials, i.e., whether two signals systematically rise and fall together (Cui et al., 2023). The frequency band of interest for investigating FC changes was the PPR band (3-4 Hz). PLV was calculated between all channels, and to gain more regional dependent measures, we calculated it region-wise by averaging the corresponding electrodes together. The regions included: Fronto-polar (Fp): Fp1, Fp2; Fronto-central (Fc): F3, F4, Fz; Fronto-temporal (Ft): F7, F8; Central (C): C3, C4, Cz; Temporal (T): T3, T4, T5, T6; Parietal (P): P3, P4, Pz; Occipital (O): O1, O2. The same 4 patients excluded from the PSD analysis (under section 2.5) due to missing recordings were also excluded from this analysis.

### 2.7 Statistical analysis

All analyses and statistical tests were carried out in Python 3.12 (Python Software Foundation). Our data showed unequal variances and was not normally distributed. Hence, for all comparisons between groups, we used robust permutation tests. Permutation tests can be applied to non-normally distributed data and control for family-wise error rates for comparisons in multidimensional data, making them well-suited for FC calculations channel- and region-wise (Smith & Nicols 2008; Rempala & Yang 2013). We used the difference between sample means as test statistics, with 1000 permutations generated with random resampling. Bonferroni correction was applied to correct for multiple comparisons between groups. All statistical tests were independent and two-sided. The level of significance was set at p < 0.05. 95 % CI was calculated for significant differences with Bootstrapping along the exact specifications as the permutation test.

## 3. Results

### 3.1 Study cohorts

A total of 84 participants were included in this analysis. Our cohort includes children and adults and is predominantly female (Table 1) since PSE is more prevalent in females, children, and adolescents (Fisher et al., 2022). Importantly, we found no significant age difference between any of our groups (all p > 1.0).

### 3.2. Visual evoked potentials (P100)

First, to characterize the general reactivity of the primary visual cortex to photic stimulation, we measured the P100. The latency of the P100 showed no significant differences between PSE patients and HC (all p > 0.119), in line with previous findings (Brinciotti et al., 2020; Siniatchkin et al., 2007b). The PWE group did, however, exhibit a significantly larger latency compared to HC (p = 0.008, CI_95_ = [-29.632, -12.067]) (Figure 1A&B). Contrary to the previous literature, the P100 (peak-to-peak) amplitude showed no significant differences between HC and either of the patient groups (PWE or PSE) (all p > 0.152) (Figure 1A&B). Overall, we observed no significant P100 amplitude or latency alterations in PSE patients compared to control subjects.

**Figure 1.:**
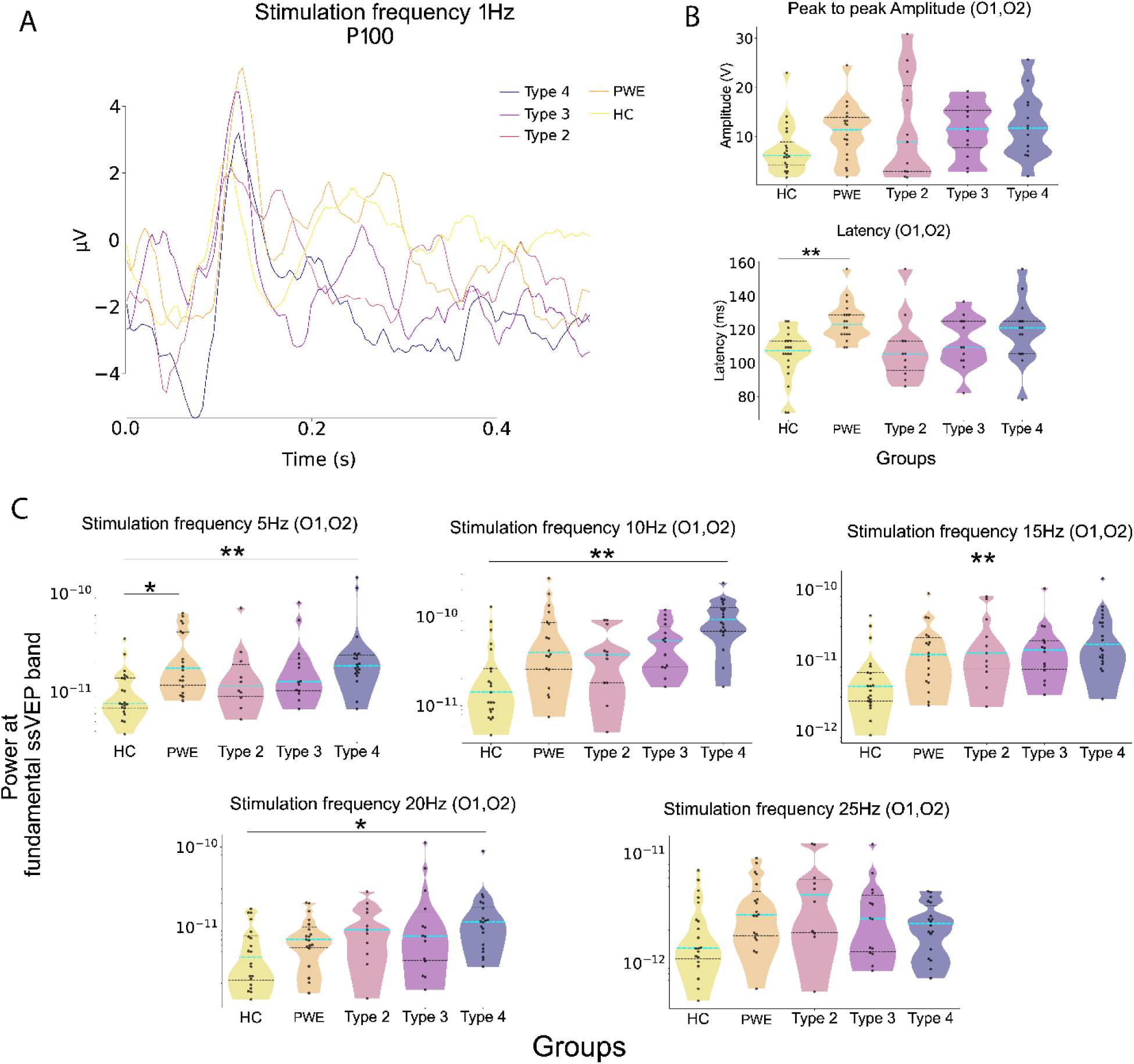
P100 and ssVEP characteristics for different stimulation frequencies. (**A**) Group averages of the P100 amplitude in response to 1 Hz stimulation frequency per group. (**B**) Violin plots show the peak-to-peak amplitude and latency of the P100 response. No significant differences in amplitude appear (top). PWE shows significantly longer latency (bottom) compared to HC. (**C**) Violin plots display the log-transformed absolute power of the fundamental ssVEP at occipital electrodes (O1, O2) under the five stimulation frequencies of interest. Significant differences compared to HC are apparent in Type 4, which shows heightened activity for almost all stimulation frequencies. PWE also showed significantly increased power compared to HC for stimulation frequency 5 Hz. Blue lines show the median, while the thinner black lines represent the lower and upper quartiles of the data. Individuals are represented as dots. Statistical significance is represented as *p ≤ 0.05, **p ≤ 0.01 and ***p ≤ 0.001.

### 3.3 Steady-state visual evoked potentials and Photoparoxysmal responses

Next, we analyzed the occipital ssVEP in response to higher temporal frequencies to further investigate the reactivity of the primary visual cortex (Porciatti et al., 2000). This was assessed at the fundamental frequency band, corresponding to the narrow band around the stimulation frequency. The fundamental ssVEP power increased significantly in the PSE Type 4 group, compared to HC for stimulation frequencies 5 Hz, 15 Hz, and 20 Hz. However, this remained unchanged in Types 3 and 2 PSE patients (Figure 1C; Supplementary Table 1). We also observed a significantly increased ssVEP power in PWE compared to HC for stimulation frequency 5 Hz (Figure 1C; Supplementary Table 1). No further differences were found when comparing PWE to HC (Figure 1C) or to any of the PSE groups (Supplementary Section 3.1). Additional comparisons between PSE patients revealed a significant ssVEP power increase for stimulation frequency of 10 Hz in Type 4 compared to the other PSE groups (Supplementary Section 3.2.).

To characterize the epileptic activity of PSE patients, we analyzed the occipital PPR power in response to the same temporal frequencies as for ssVEP. As expected, the PPR power increased significantly in all PSE patients compared to HC (Figure 2A; Supplementary Table 1), with the highest values observed for stimulation frequencies between 10-20 Hz (Figure 2B). The peak group power within the established 10-20 Hz risk range was noted for different stimulation frequencies (15 Hz for Type 2 and 3; and 10 Hz for Type 4 patients). PPR power increased with type severity, showing the highest values in Type 4 followed by Type 3 (Figure 2A&B, Table 2). The effect of type severity was further evidenced by Type 4 reaching significantly larger occipital PPR power compared to the other PSE groups for stimulation frequencies 10-25 Hz (Supplementary Section 4.1.). For the same stimulation frequencies, Type 4 also showed significantly higher occipital PPR power compared to PWE (Supplementary Section 4.2.). Surprisingly, PWE exhibited significantly higher PPR power compared to HC for stimulation frequencies 20 Hz and 25 Hz (Figure 2A; Supplementary Table 1). However, we do not consider this an actual PPR activity since PWE showed a rise in power unspecific to a frequency band (between 1-15Hz), concurrent with the HC group showing a slight decline in power in response to these stimulation frequencies. Together, these most likely facilitated the significant difference. Lastly, all the above-described occipital PPR power patterns similarly emerged at parietal and frontal electrodes (Supplementary Sections 5 & 6).

**Figure 2.:**
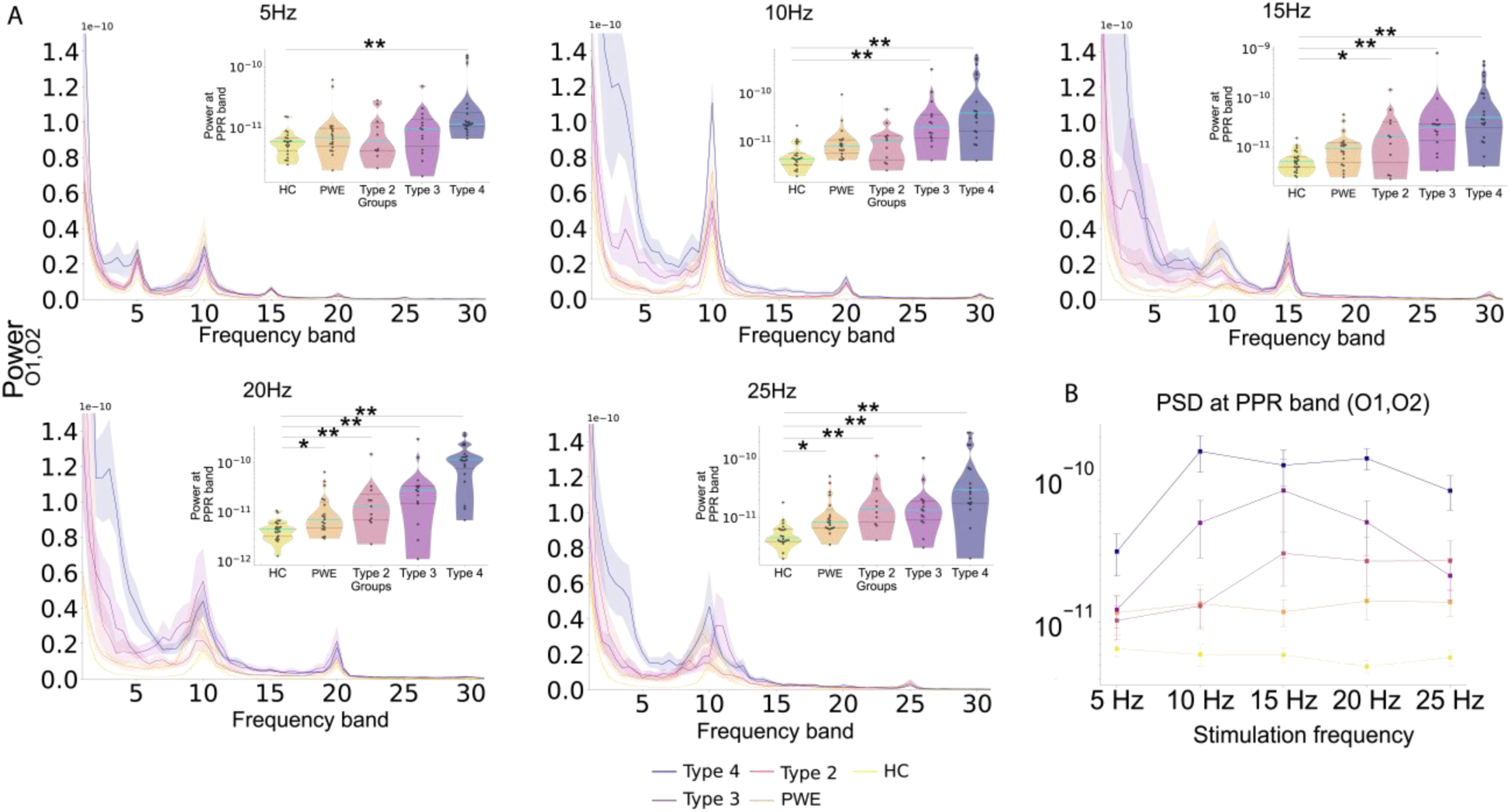
Power differences. (**A**) Power spectral densities (PSDs) displayed at each stimulation frequency averaged per group at occipital electrodes (O1, O2). PSDs show power fluctuations at the bands linked to the triggering frequency and the pathological PPR activity between 3-4 Hz (PPR band) of PSE patients. The shaded area shows the standard error corresponding to each group’s average. Stimulation frequency-specific PPRs are further displayed on the violin plots, showing a gradual increase with type severity for stimulation frequencies 10, 15, 20, and 25 Hz. Blue lines show the median while the thinner black lines represent the lower and upper quartiles of the data. Individuals are represented as dots. (**B**) The line plot displays the average power, with standard error, for all. It shows an increase in PPR activity for at stimulation frequencies that were previously shown to be high risk (10, 15, and 20 Hz) and a still high but decreasing PPR for 25 Hz. Both the violin plots and the line plot show log-transformed values.

In summary, Type 4 patients showed increased occipital ssVEP power at the fundamental frequency band compared to HC but revealed no difference when measured against PWE. Spectral power in the PPR band exhibited an increase governed by type severity at the occipital, as well as parietal and frontal electrodes. The highest PPR power appeared in Type 3 and 4 PSE groups, specifically for stimulation frequencies 10 - 20 Hz (Figure 2A & B).

### 3.4 Scalp-scalp Connectivity

Finally, functional network effects were explored by analyzing FC patterns in the 3-4Hz frequency band (PPR band) in response to stimulation frequencies 5-25 Hz. We identified only a few decreased FC (hypoconnected) patterns in PWE compared to HC. These hypoconnected FC patterns were found between central and frontotemporal regions for stimulation frequencies 5 and 20 Hz and between temporal and occipital regions for stimulation frequencies 10 Hz (Figure 3). In contrast, different FC patterns in PSE patients compared to HC were apparent for risk range frequencies (10-20 Hz) established in our PSD analysis. We observed significant hyper- and hypoconnectivity patterns in PSE patients compared to HC, governed by type severity and stimulation frequency. The most prominent patterns of decreased FC in all PSE patients were shown for stimulation frequencies 10-25 Hz between central and parietal regions and between central and frontopolar regions (Figure 3; Supplementary Table 2). This centro-parietal and centro-frontopolar hypoconnectivity was less pronounced when comparing PSE patients to PWE (Supplementary Section 7 & Figure 3). Increased FC (hyperconnectivity) patterns were only observed in patients with PPR propagation, i.e., in Types 3 and 4. These were subtle in Type 3, with a slight FC increase between frontal electrodes and between occipital and frontal channels (Figure 3). Hyperconnectivity became quite prominent in Type 4, specifically between frontocentral and frontopolar and between occipital and frontocentral regions for stimulation frequencies 10-20 Hz (Figure 3; Supplementary Table 3). Significantly increased within-anterior and between-posterior-anterior FC seemed to be even more evident in Types 3 and 4 compared to PWE (Supplementary Section 7 & Figure 3).

**Figure 3:**
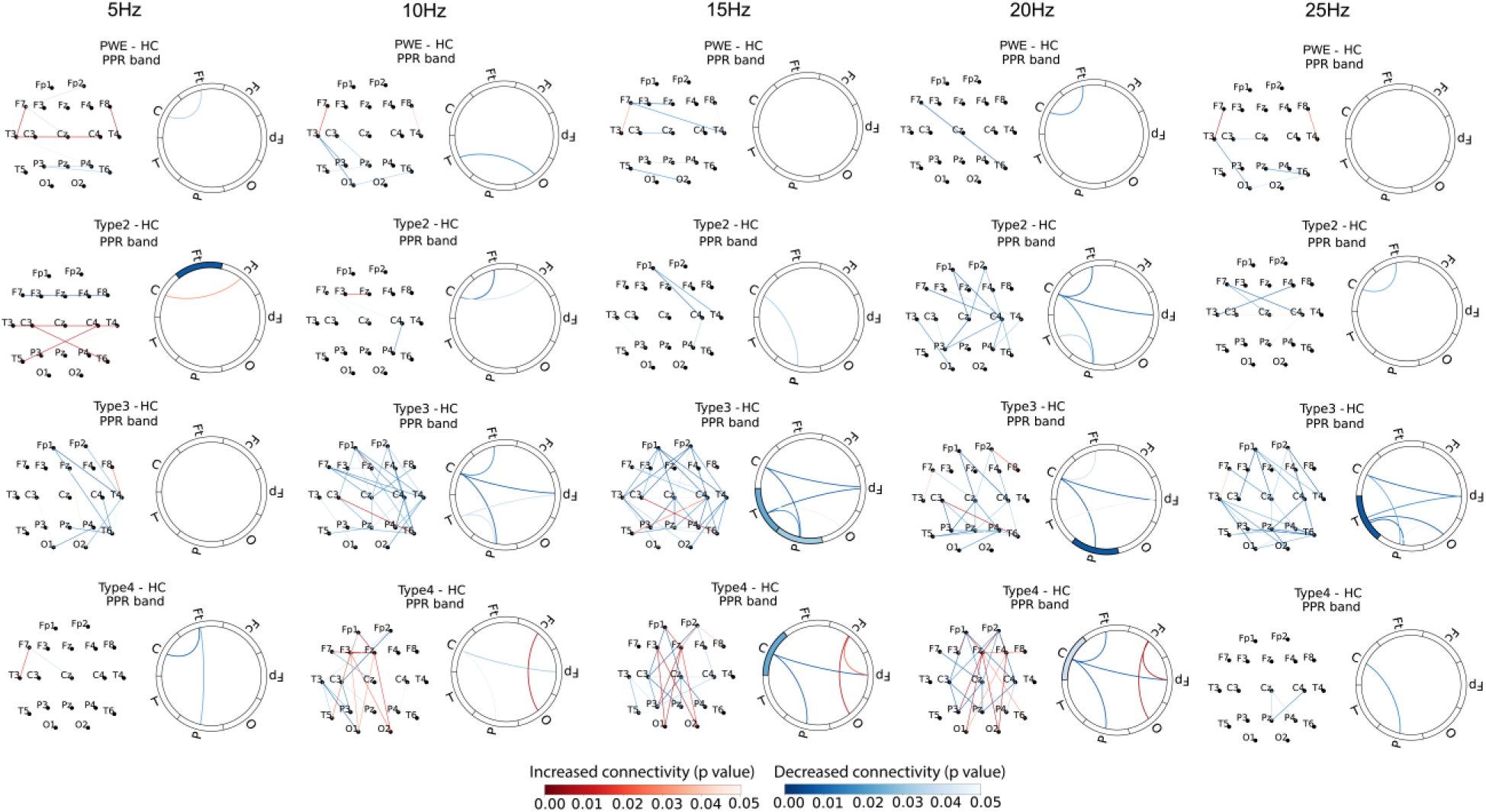
FC patterns in comparison to HC. Significant FC patterns are shown channel (topo plots) and region-wise (spider plots) for all five stimulation frequencies. The red color indicates increased, while the blue color indicates decreased connectivity in patients compared to HC. This was calculated by subtracting HC from the respective patient group (PWE, 2, 3, and 4). The shade of the colors corresponds to the level of significance, with darker colors indicating higher significance. In the case of a significant within-region connectivity difference, the level of significance and the nature of change (decrease, increase) are indicated by the colored in sections at the corresponding region of the spider plot. Hyperconnectivity shows an increase with type severity between occipital and frontal and within frontal regions, mainly in the case of Type 4, but some patterns are noticeable in Type 3 as well. Hypoconnectivity is present between central and parietal and central and frontopolar regions in all PSE types compared to HC.

In brief, we showed FC differences in PSE patients’ responses to IPS compared to control subjects. These involved hypoconnected centro-parietal and centro-frontopolar areas in all PSE types and hyperconnected frontal occipito-frontal connectivity in PSE patients with PPR propagation.

## 4. Discussion

We examined the underlying mechanisms of type-specific PSE responses using IPS and scalp EEG, aiming to provide a comprehensive, systematic analysis of PSE and reveal new insights into the underlying mechanisms of the condition. To date, multiple studies have investigated PSE and the hyperexcitability of the visual cortex (Brinciotti et al., 2020; Siniatchkin et al., 2007a,b; Strigaro et al., 2012; Wilkins et al., 2004). However, possible network differences driving the propagation of the PPR have yet to be investigated in detail. In this present study, we observed that PPR activity was affected by both type severity and stimulation frequency. However, hyperexcitability of the visual cortex was only apparent in Type 4 patients, posing the question to what extent this can be responsible for the generation and, more importantly, the propagation of the PPR. The altered FC profiles in Type 3 and Type 4 PSE patients suggest that large-scale network alterations could underlie PSE and the propagation of the PPR.

Our results show evident pathological activity (PPR) at occipital electrodes in PSE patients, with significantly enhanced PPR EEG power compared to HC. The occipital PPR power was affected by stimulation frequency, with the highest values shown for the range of 10-20 Hz, in line with previous studies, and by type severity, with Type 4 showing the highest PPR power (Harding & Harding, 2010; Kasteleijn-Nolst Trenité et al., 2012; Porciatti et al., 2000; Taoufiqi et al., 2016; Zhang et al., 2023). As part of a supplementary analysis, we identified similar patterns at parietal and frontal channels, which further solidifies the involvement of these two areas in PPR propagation, as previously suggested by Moeller et al. (2013; 2009a).

Unlike prior studies, we did not observe an increased P100 amplitude in PSE patients, which was previously proposed as an indication of the hyperexcitability of the visual cortex and key to the PSE reaction. (Brinciotti et al., 2020; Genç et al., 2005; Siniatchkin et al., 2007b). Albeit not significant, we showed trends for the highest P100 amplitudes in Type 4 and lowest in the HC group, similar to previous findings using single flashes (Strigaro et al., 2012). Interestingly, a trend for increased P100 amplitude also arose in PWE compared to HC. Although PWE individuals are not photosensitive, their responses could deviate from those of the HC due to underlying neurological differences. The latency of the P100 only showed a significant increase in PWE compared to HC, which is not unprecedented (Genç et al., 2005). Several factors can account for prolonged latency in these patients, such as ASMs (Geller et al., 2005; Verrotti et al., 2000), abnormal synaptic transmission (Gokcay et al., 2003; Mervaala et al., 1987), or minor changes in underlying neurophysiologic structures (microdysgenesia) (Meencke & Janz, 1985), many of which could also apply to the PWE group.

The occipital ssVEP power, however, did increase significantly at the fundamental frequency band in Type 4 compared to HC in line with Poricatti et al. (2000). This provides evidence that PSE patients have heightened occipital activity in response to ongoing visual stimulation, which we only found a trend for in the P100 analysis. The fact that ssVEP power only increased in Type 4 compared to HC agrees with the study of Siniatchkin et al. (2007b), which observed higher VEP amplitudes in patients with propagating PPRs compared to patients with more local PPRs. The ssVEP power also increased in PWE compared to HC, although for only stimulation frequency 5 Hz. Such a ssVEP power increase argues for increased neuronal excitability in the visual cortex of not only the PSE but also non-PSE epilepsy patients compared to HC. The absence of a significant ssVEP power difference between PWE and the PSE groups further supports this claim. Together, these findings suggest that PSE patients do show hyperexcitability in the visual cortex to external visual stimulation, also shown by previous papers (Brinciotti et al., 2020b; Moeller et al., 2009a; Siniatchkin et al., 2007b; Strigaro et al., 2012). However, in our study, this local effect was small and not significantly different from those in PWE. Hence, a hyperexcitable visual cortex might be a requirement for PPR, however, it most likely must be coupled with other factors to generate and propagate the pathological response of PSE patients. This aligns with a study by Moeller et al. (2009a), which showed that in PSE patients, PPR was associated with activity in frontal and parietal regions, suggesting that the PPR depends on network differences rather than overactivity of the occipital cortex alone. Another study found higher visual cortex activation in HC compared to PSE patients during IPS, which could support the notion that a hyperexcited visual cortex in itself is insufficient to trigger pathological activity (Bartolini et al., 2014), and instead, it might depend on functional network dynamics (Varotto et al., 2012).

Our FC results corroborate this hypothesis, showing significantly different connectivity patterns observed in PSE patients compared to HC and to PWE in response to stimulation at high-risk frequencies (10-20 Hz). We argue that these pathological hypo-and hyperconnectivity alterations drive the PPR and are responsible for its propagation.

All PSE patients demonstrated hypoconnectivity between central and parietal, central and frontopolar, and central and frontotemporal regions when compared to HC. We infer that these subnetworks likely correspond to the default mode network (DMN), a collection of regions active when the brain is at rest and not engaged in an activity. It includes the frontal central and parietal areas with the ventral and dorsal medial prefrontal cortex, posterior cingulate cortex, and lateral parietal cortex (Raichle, 2015). The DMN was previously shown to decrease its activity in PSE patients and to further decrease during PPR (Bartolini et al., 2014; Moeller, et al., 2009b). Resting-state DMN hypoconnectivity was also shown to accompany multiple epilepsy syndromes (Gonen et al., 2020; Luo et al., 2011; McGill et al., 2012; Parsons et al., 2020). Hence, it is not surprising that PWE shows some minimal hypoconnectivity between DMN-related areas compared to HC. The fact that PWE also exhibited some hypoconnectivity in the DMN could account for less pronounced hypoconnectivity differences when compared to PSE patients. Furthermore, decreased connectivity between DMN regions was suggested to facilitate hyperconnectivity between other areas (Vollmar et al., 2012), which could fit our observations of several hyperconnected electrodes with concurrent hypoconnectivity in the inferred DMN regions (Figure 3).

As the severity of the PSE type increased, hyperconnectivity became more evident, demonstrated by increasingly widespread patterns compared to HC. While hypoconnectivity was observed in all PSE patients, hyperconnectivity only characterized those with PPR propagation in response to the high-risk stimulation frequencies. Type 3 patients showed some abnormal connectivity of the posterior regions and a slight increase in FC between occipital and frontal electrodes and between frontal electrodes. Type 4 patients with generalized PPR had quite a clear posterior-anterior connectivity, the directionality of which was modeled in a study by Moeller et al. (2013), and within anterior hyperconnectivity. The increased anterior connectivity agrees with findings from Varotto et al. (2012) who found hyperconnectivity within anterior regions of PSE patients compared to HC. Increased synchronization of frontoparietal networks was also associated with PPR in a study on baboons (Szabo et al., 2016). In a supplementary analysis, hyperconnectivity patterns in Type 3 and 4 seemed to be enhanced compared to PWE, as opposed to when these groups were contrasted with HC, particularly in Type 3, where hyperconnectivity was much more evident. Our ssVEP analysis indicated that, in general, epilepsy patients might have higher local susceptibility to visual stimuli independent from PSE. It could be that, as a protective mechanism, in PWE, these areas (anterior regions and posterior-anterior regions) are less connected to prevent a PPR from occurring.

### 4.1.1 Limitations and outlook

There is a possibility that the lack of a significant P100 amplitude increase in all patient groups compared to HC may be due to ASM intake. However, this remains debatable, as some studies have reported reduced VEP amplitude in patients taking ASM (Geller et al., 2005; Kanazawa & Nagafuji, 1997), while others claim that it only affects the latency rather than the amplitude of the P100 (Verrotti et al., 2000; Yüksel et al., 1995). Another limitation we must address is the sex imbalance between the HC group (mostly male) and the patient groups (mostly female). It has been shown that generally, females have higher ssVEP amplitudes compared to males (Krishnan et al., 2005; Skosnik et al., 2006). In this present study, we cannot entirely exclude whether the significant ssVEP power increase in patient groups was only due to sex differences or due to a combination of sex and photosensitivity effects. However, we consider the former unlikely since we observed variability in the ssVEP power governed by stimulation frequency and type severity, as evidenced by only Type 4 patients showing significant differences compared to HC. Regarding FC analyses, we noted minimal distinctions between PWE and HC. Since PWE and PSE groups are similar in sex distribution, this indicates that the variations in FC between PSE patients and HC were predominantly influenced by PSE type, the presence of PPR, and the stimulation frequency rather than sex differences. Finally, in the present study, the analyzed FC patterns were restricted only to responses to periodically flashing lights. In the future, functional network alterations could also be studied in reaction to a variety of visual triggers that better represent real-life environments, e.g., stimuli of periodically varying chromaticity or stimuli of differing field size.

## 5. Conclusions

Overall, the results of our retrospective study suggest that the visual cortex of PSE patients is hyperexcitable and remains significantly more active during higher stimulation frequencies of the IPS compared to HC. Furthermore, we showed that PSE-type severity increases the magnitude of the PPR at occipital, parietal, and frontal electrodes. However, the hyperexcitability of the cortex is not the only determinant of PPR and PSE in general, as we did not observe increased activity in the occipital region of non-PSE patients compared to PSE patients. Instead, it appears that PPR relies on a hyperexcitable visual cortex, providing the basis for a local frequency-specific cortical overactivation and an altered brain network connectivity that propagates this overactivation in a type-specific manner.

## Data availability

Data and code for analysis will be made available upon reasonable request.

