## Supplementary material for "Functional network dynamics in photosensitive epilepsy depend on stimulation frequency and photosensitivity type"

#### 1. Statistical comparisons of the ssVEP and PPR power

| Stimulation frequency | Comparisons | P-values (ssVEP) | Confidence intervals ($CI_{95}$) (ssVEP) | P-values (PPR) | Confidence intervals ($CI_{95}$) (PPR) |
|---|---|---|---|---|---|
| 5 Hz | PWE – HC | **0.016** | **[-2.453$_{e-11}$, -6.885$_{e-12}$]** | 0.679 | [-1.791$_{e-11}$, -3.515 $_{e-13}$] |
| | Type 2 – HC | 0.783 | [-2.578$_{e-11}$, 5.763$_{e-13}$] | 0.495 | [-1.036$_{e-11}$, 7.719$_{e-13}$] |
| | Type 3 – HC | 0.159 | [-2.592$_{e-11}$, -3.184$_{e-12}$] | 0.088 | [-1.499$_{e-11}$, -1.578$_{e-12}$] |
| | Type 4 – HC | **0.008** | **[-4.163$_{e-11}$, -7.961$_{e-12}$]** | **0.008** | **[-5.310$_{e-11}$, -1.041$_{e-11}$]** |
| 10 Hz | PWE – HC | 0.096 | [-8.053$_{e-11}$, -1.438$_{e-11}$] | 0.128 | [-2.610$_{e-11}$, -2.091$_{e-12}$] |
| | Type 2 – HC | 0.999 | [-3.842$_{e-11}$, 6.926$_{e-12}$] | 0.176 | [-1.844$_{e-11}$, -1.351$_{e-12}$] |
| | Type 3 – HC | 0.064 | [-4.944$_{e-11}$, -7.559$_{e-12}$] | **0.008** | **[-1.142$_{e-10}$, -1.795$_{e-11}$]** |
| | Type 4 – HC | **0.008** | **[-9.805$_{e-11}$, -4.744$_{e-11}$]** | **0.008** | **[-2.566$_{e-10}$, -7.754$_{e-11}$]** |
| 15 Hz | PWE – HC | 0.112 | [-2.355$_{e-11}$, -2.485$_{e-12}$] | 0.096 | [-1.428$_{e-11}$, -2.445$_{e-12}$] |
| | Type 2 – HC | 0.096 | [-3.899$_{e-11}$, -4.816$_{e-12}$] | **0.024** | **[-6.262$_{e-11}$, -7.700$_{e-12}$]** |
| | Type 3 – HC | 0.199 | [-3.545$_{e-11}$, -3.370$_{e-12}$] | **0.008** | **[-3.088$_{e-10}$, -1.511$_{e-11}$]** |
| | Type 4 – HC | **0.008** | **[-4.412$_{e-11}$, -1.084$_{e-11}$]** | **0.008** | **[-2.153$_{e-10}$, -6.508$_{e-11}$]** |
| 20 Hz | PWE – HC | 0.671 | [-5.411$_{e-12}$, 1.175$_{e-12}$] | **0.032** | **[-2.010$_{e-11}$, -3.880$_{e-12}$]** |
| | Type 2 – HC | 0.184 | [-1.069$_{e-11}$, -7.904$_{e-13}$] | **0.008** | **[-6.309$_{e-11}$, -6.602$_{e-12}$]** |
| | Type 3 – HC | 0.159 | [-4.147$_{e-11}$, -2.481$_{e-12}$] | **0.008** | **[-1.148$_{e-10}$, -1.808$_{e-11}$]** |
| | Type 4 – HC | **0.016** | **[-2.130$_{e-11}$, -4.041$_{e-12}$]** | **0.008** | **[-1.932$_{e-10}$, -9.903$_{e-11}$]** |
| 25 Hz | PWE – HC | 0.279 | [-2.859$_{e-12}$, -1.928$_{e-13}$] | **0.024** | **[-1.629$_{e-11}$, -3.779$_{e-12}$]** |
| | Type 2 – HC | 0.079 | [-5.579$_{e-12}$, -6.578$_{e-13}$] | **0.008** | **[-5.010$_{e-11}$, -7.624$_{e-12}$]** |
| | Type 3 – HC | 0.879 | [-3.205$_{e-12}$, 4.235$_{e-13}$] | **0.008** | **[-3.538$_{e-11}$, -6.785$_{e-12}$]** |
| | Type 4 – HC | >1.0 | [-9.558$_{e-13}$, 9.138$_{e-13}$] | **0.008** | **[-1.275$_{e-10}$, -3.622$_{e-11}$]** |

**Table 1.:** Statistical between-group comparisons of the ssVEP and PPR power per stimulation frequency. Significant differences are shown in bold. ssVEP = Steady-state evoked potential; PPR = Photoparoxysmal response; HC= Healthy controls; PWE = Patients with epilepsy

### 2. Statistical comparisons of PLV values

| Stimulation frequency | Comparisons | P-values (C-P) | Confidence intervals ($CI_{95}$) (C-P) | P-values (C-Fp) | Confidence intervals ($CI_{95}$) (C-Fp) |
|---|---|---|---|---|---|
| 5 Hz | PWE – HC | 0.359 | [-0.011, 0.162] | 0.272 | [-0.001, 0.169] |
| | Type 2 – HC | 0.575 | [-0.015, 0.151] | 0.855 | [-0.001, 0.164] |
| | Type 3 – HC | 0.072 | [-0.006, 0.154] | 0.088 | [0.004, 0.164] |
| | Type 4 – HC | 0.247 | [-0.008, 0.156] | 0.751 | [-0.005, 0.155] |
| 10 Hz | PWE – HC | 0.495 | [-0.015, 0.128] | >1.0 | [-0.027, 0,124] |
| | Type 2 – HC | 0.056 | [-0.015, 0.126] | 0.743 | [-0.033, 0.124] |
| | Type 3 – HC | **0.008** | **[-1.200$_{e-02}$, 0.126]** | **0.008** | **[-3.436$_{e-02}$, 0.118]** |
| | Type 4 – HC | **0.048** | **[-0.009, 0.127]** | **0.032** | **[-0.029, 0.126]** |
| 15 Hz | PWE – HC | 0.799 | [-0.028, 0.131] | >1.0 | [-0.049, 0.116] |
| | Type 2 – HC | **0.032** | **[-0.021, 0.132]** | 0.064 | [-0.041, 0.122] |
| | Type 3 – HC | **0.008** | **[-0.021, 0.137]** | **0.008** | **[-0.047, 0.119]** |
| | Type 4 – HC | **0.008** | **[-0.029, 0.123]** | **0.008** | **[-0.046, 0.119]** |
| 20 Hz | PWE – HC | 0.279 | [-0.004, 0.136] | 0.159 | [0.011, 0.157] |
| | Type 2 – HC | **0.016** | **[-7.653$_{e-04}$, 0.136]** | **0.008** | **[9.237$_{e-03}$, 0.151]** |
| | Type 3 – HC | **0.008** | **[-0.002, 0.142]** | **0.008** | **[0.014, 0.160]** |
| | Type 4 – HC | **0.008** | **[-0.003, 0.142]** | **0.008** | **[0.007, 0.163]** |
| 25 Hz | PWE – HC | >1.0 | [-0.039, 0.107] | 0.775 | [-0.029, 0.114] |
| | Type 2 – HC | 0.264 | [-0.037, 0.108] | 0.159 | [-0.029, 0.121] |
| | Type 3 – HC | **0.024** | **[-0.039, 0.101]** | **0.008** | **[-0.031, 0.117]** |
| | Type 4 – HC | **0.016** | **[-0.039, 0.107]** | 0.152 | [-0.030, 0.118] |

**Table 2.** Statistical between-group comparisons of the hypoconnected regions (PLV values) at the PPR band (3-4 Hz) per stimulation frequency. Significant differences are shown in bold. PPR = Photoparoxysmal response; C = Central; P = Parietal; Fp = Frontopolar; HC = Healthy controls; PWE = Patients with epilepsy.

| Stimulation frequency | Comparisons | P-values (Fp-Fc) | Confidence intervals ($CI_{95}$) (Fp-Fc) | P-values (Fc-O) | Confidence intervals ($CI_{95}$) (Fc-O) |
|---|---|---|---|---|---|
| 5 Hz | PWE – HC | >1.0 | [-0.075, 0.107] | >1.0 | [-0.080, 0.051] |
| | Type 2 – HC | >1.0 | [-0.081, 0.101] | >1.0 | [-0.078, 0.050] |
| | Type 3 – HC | 0.967 | [-0.085, 0.100] | 0.088 | [-0.075, 0.055] |
| | Type 4 – HC | >1.0 | [-0.088, 0.095] | 0.119 | [-0.085, 0.050] |
| 10 Hz | PWE – HC | >1.0 | [-0.092, 0.122] | >1.0 | [-0.068,0.103] |
| | Type 2 – HC | >1.0 | [-0.096, 0.113] | >1.0 | [-0.084, 0.093] |
| | Type 3 – HC | 0.336 | [-1.002$_{e-01}$, 0.106] | 0.392 | [-6.505$_{e-02}$, 0.105] |
| | Type 4 – HC | 0.088 | [-0.092, 0.118] | **0.008** | **[-0.070, 0.098]** |
| 15 Hz | PWE – HC | >1.0 | [-0.115, 0.084] | >1.0 | [-0.084, 0.061] |
| | Type 2 – HC | >1.0 | [-0.119, 0.086] | 0.224 | [-0.084, 0.060] |
| | Type 3 – HC | 0.112 | [-0.127, 0.095] | 0.096 | [-0.099, 0.058] |
| | Type 4 – HC | **0.024** | **[-0.120, 0.088]** | **0.008** | **[-0.081, 0.057]** |
| 20 Hz | PWE – HC | >1.0 | [-0.135, 0.078] | >1.0 | [-0.079, 0.065] |
| | Type 2 – HC | >1.0 | [-1.261$_{e-01}$, 0.075] | 0.759 | [-8.599$_{e-02}$, 0.055] |
| | Type 3 – HC | 0.224 | [-0.124, 0.087] | 0.855 | [-0.082, 0.059] |
| | Type 4 – HC | **0.008** | **[-0.138, 0.068]** | **0.008** | **[-0.080, 0.063]** |
| 25 Hz | PWE – HC | >1.0 | [-0.074, 0.129] | >1.0 | [-0.048, 0.117] |
| | Type 2 – HC | >1.0 | [-0.083, 0.117] | >1.0 | [-0.058, 0.108] |
| | Type 3 – HC | >1.0 | [-0.077, 0.118] | >1.0 | [-0.054, 0.109] |
| | Type 4 – HC | >1.0 | [-0.075, 0.126] | 0.839 | [-0.049, 0.117] |

**Table 3.** Statistical between-group comparisons of the hyperconnected regions (PLV values) at the PPR band (3-4 Hz) per stimulation frequency. Significant differences are shown in bold. PPR = Photoparoxysmal response; Fc= Frontocentral; Fp = Frontopolar; O = Occipital; HC = Healthy controls; PWE = Patients with epilepsy.

### 3. ssVEP power at Occipital channels

#### 3.1. Occipital ssVEP power in comparison to PWE

In our main analysis, we compared all five groups along the elicited ssVEP power at the fundamental occipital ssVEP in response to the five stimulation frequencies of interest. Here, we investigated how PSE groups differ from PWE. Pairwise comparisons at occipital electrodes revealed no significant differences in PWE compared to any of the PSE patients for neither of the five stimulation frequencies (5 Hz: $p > 1.0$.; 10 Hz: $p > 0.419$; 15 Hz: $p > 0.857$; 20Hz $p > 0.149$; 25Hz $p > 0.227$).

#### 3.2. Occipital ssVEP power in comparison Type 4

Next, we compared PSE groups (Type 2 and 3) to Type 4. Occipital ssVEP power at the fundamental frequency band showed significantly higher ssVEP power in Type 4 compared to Type 2 ($p = 0.004$, $CI_{95} = [-8.768_{e-11}, -2.732_{e-11}]$) and Type 3 patients ($p = 0.019$, $CI_{95} = [-7.332_{e-11}, -1.792_{e-11}]$) for stimulation frequency 10 Hz. All other comparisons at the remaining stimulation frequencies were non-significant (5 Hz: $p > 0.935$; 15 Hz: $p > 0.986$; 20 Hz $p > 1.0$; 25 Hz $p > 0.059$). All reported p-values are Bonferroni corrected.

### 4. PPR power at Occipital channels

#### 4.1. Occipital PPR power in comparison to Type 4

We compared the PPR power of PSE groups to Type 4 at occipital electrodes. At the PPR frequency band, as expected, we observed significantly higher power in the Type 4 group compared to the PWE and some of the other PSE groups (Type 2 and 3). Pairwise comparisons revealed significantly increased PPR power in Type 4 for stimulation frequencies 10 Hz (Type 2 – Type 4: $p = 0.004$, $CI_{95} = [-2.522_{e-10}, -7.862_{e-11}]$), 20 Hz (Type 2 – Type 4 : $p = 0.004$, $CI_{95} = [-1.686_{e-10}, -6.298_{e-11}]$; Type 3 – Type 4: $p = 0.012$, $CI_{95} = [-1.534_{e-10}, -1.860_{e-11}]$), and 25 Hz (Type 3 – Type 4: $p = 0.039$, $CI_{95} = [-1.168_{e-10}, -2.419_{e-11}]$). For stimulation frequency 5 Hz, all p-values > 0.304, and for stimulation frequency 15 Hz, all p-values> 0.116.

#### 4.2. Occipital PPR power in comparison PWE

Next, we investigated how PSE groups differ from PWE at the PPR frequency band. As expected, we observed significantly higher PPR power in the Type 4 group compared to the PWE for stimulation frequencies 10 Hz (PWE – Type 4: $p = 0.006$, $CI_{95} = [-2.618_{e-10}, -7.261_{e-11}]$), 15 Hz (PWE – Type 4: $p = 0.006$, $CI_{95} = [-2.036_{e-10}, -5.779_{e-11}]$), 20 Hz (PWE – Type 4 : $p = 0.006$, $CI_{95} = [-1.869_{e-10}, -8.772_{e-11}]$), and 25 Hz (PWE – Type 4: $p = 0.006$, $CI_{95} = [-1.262_{e-10}, -3.410_{e-11}]$). For stimulation frequency 5Hz, all p-values > 0.444. All reported p-values are Bonferroni corrected.

### 5. PPR power Parietal channels

#### 5.1. Parietal PPR power in comparison to HC

To complement our main analysis, we calculated the power spectral properties of parietal and frontal channels at the PPR frequency band. For parietal channels, this included P3, P4, and Pz electrodes, while for frontal channels, F3, F4, and Fz were used. For methods and calculation of spectral properties, refer to our main text's methods and materials section.

Pairwise comparisons of the parietal PPR revealed a significant increase in PSE patients compared to HC for stimulation frequencies 5 Hz (Type 3 – HC: $p = 0.024$, $CI_{95} = [-2.106_{e-11}, -1.788_{e-12}]$; Type 4 – HC: $p = 0.008$, $CI_{95} = [-2.833_{e-11}, -7.166_{e-12}]$), 10 Hz (Type 3 – HC: $p = 0.008$, $CI_{95} = [-3.132_{e-11}, -8.709_{e-12}]$; Type 4 – HC: $p = 0.008$ , $CI_{95} = [-1.462_{e-10}, -4.065_{e-11}]$), 15 Hz (Type 2 – HC: $p = 0.039$, $CI_{95} = [-2.203_{e-11}, -3.126_{e-12}]$; Type 3 – HC: $p = 0.008$ , $CI_{95} = [-1.567_{e-10}, -9.079_{e-12}]$; Type 4 – HC: $p = 0.008$ , $CI_{95} = [-1.529_{e-10}, -3.988_{e-11}]$), 20 Hz (Type2 – HC: $p = 0.008$, $CI_{95} = [-2.559_{e-11}, -3.798_{e-12}]$; Type 3 – HC: $p = 0.008$, $CI_{95} = [-1.014_{e-10}, -1.259_{e-11}]$; Type 4 – HC: $p = 0.008$, $CI_{95} = [-1.298_{e-10}, -5.462_{e-11}]$), and 25 Hz (Type2 – HC: $p = 0.008$, $CI_{95} = [-2.503_{e-11}, -3.539_{e-12}]$; Type 3 – HC: $p = 0.008$, $CI_{95} = [-2.379_{e-11}, -4.646_{e-12}]$; Type 4 – HC: $p = 0.008$, $CI_{95} = [-5.510_{e-11}, -2.075_{e-11}]$). For stimulation frequencies 20 Hz and 25 Hz we observed a significant power increase in PWE compared to HC (20 Hz: PWE – HC: $p = 0.008$, $CI_{95} = [-1.397_{e-11}, -2.532_{e-12}]$, 25Hz: PWE – HC: $p = 0.016$, $CI_{95} = [-1.239_{e-11}, -2.881_{e-12}]$), similar to what we have found at occipital electrodes. All comparisons are shown in Figure 1A&B.

**5.2. Parietal PPR power in comparison to PWE**

Additionally, we investigated parietal PPR power differences in PSE groups compared to PWE for all stimulation frequencies. Pairwise comparisons showed significantly higher PPR power in Type 4 compared to PWE for stimulation frequencies 10 Hz ($p = 0.006$, $CI_{95} = [-1.439_{e-10}, -4.018_{e-11}]$), 15 Hz ($p = 0.006$, $CI_{95} = [-1.581_{e-10}, -3.643_{e-11}]$), 20 Hz ($p = 0.006$, $CI_{95} = [-1.323_{e-10}, -5.145_{e-11}]$) and 25 Hz ($p = 0.012$, $CI_{95} = [-4.929_{e-11}, -1.378_{e-11}]$). For stimulation frequency 5 Hz, all $p > 0.257$. Type 2 and Type 3 groups showed no significant difference from PWE parietal PPR power.

**5.3. Parietal PPR power in comparison to Type 4**

Finally, we compared PSE groups (Type 2 and 3) to Type 4. Pairwise comparisons showed a significant increase of PPR power in Type 4 compared to the other PSE groups for stimulation frequencies 10 Hz (Type 2 – Type 4: $p = 0.004$, $CI_{95} = [-1.436_{e-10}, -4.031_{e-11}]$), 15 Hz (Type 2 – Type 4: $p = 0.016$, $CI_{95} = [-1.675_{e-10}, -3.185_{e-11}]$), 20Hz (Type 2 – Type 4: $p = 0.004$, $CI_{95} = [-1.166_{e-10}, -4.300_{e-11}]$), and 25Hz (Type 3 – Type 4: $p = 0.048$, $CI_{95} = [-4.367_{e-11}, -5.991_{e-12}]$). For stimulation frequency 5 Hz, all $p > 0.252$. All reported p-values are Bonferroni corrected.

### 6. PPR power at Frontal channels

**6.1. Frontal PPR power in comparison to HC**

The same line of analysis as for parietal and occipital electrodes was performed now at frontal electrodes (F3, F4, Fz). We found that the frontal PPR power significantly increased in PSE patients compared to HC for stimulation frequencies 5 Hz (Type 3 – HC: $p = 0.008$, $CI_{95} = [-8.743_{e-12}, -1.496_{e-12}]$; Type 4 – HC; $p = 0.008$, $CI_{95} = [-6.896_{e-11}, -5.713_{e-12}]$), 10 Hz (Type 3 – HC: $p = 0.008$, $CI_{95} = [-8.686_{e-11}, -1.494_{e-11}]$; Type 4 – HC; $p = 0.008$, $CI_{95} = [-3.222_{e-10}, -9.542_{e-11}]$), 15 Hz (Type 2 – HC: $p = 0.016$ , $CI_{95} = [-1.422_{e-11}, -2.333_{e-12}]$; Type 3 – HC: $p = 0.008$, $CI_{95} = [-1.863_{e-10}, -1.093_{e-11}]$; Type 4 – HC: $p = 0.008$, $CI_{95} = [-2.398_{e-10}, -5.796_{e-11}]$),20 Hz (Type2 – HC: $p = 0.016$, $CI_{95} = [-1.087_{e-11}, -1.841_{e-12}]$; Type 3 – HC: $p = 0.008$, $CI_{95} = [-5.384_{e-11}, -1.109_{e-11}]$; Type 4 – HC: $p = 0.008$, $CI_{95} = [-2.319_{e-10}, -1.092_{e-10}]$),and 25 Hz (Type2 – HC: $p = 0.008$, $CI_{95} = [-1.042_{e-11}, -1.889_{e-12}]$; Type 3 – HC: $p = 0.008$, $CI_{95} = [-2.294_{e-11}, -4.866_{e-12}]$; Type 4 – HC: $p = 0.008$, $CI_{95} = [-2.459_{e-10}, -4.415_{e-11}]$). For stimulation frequencies 20 Hz and 25 Hz we observed a significant power increase in PWE compared to HC (20 Hz: PWE – HC: $p = 0.039$, $CI_{95} = [-1.882_{e-11}, -1.479_{e-12}]$, 25 Hz: PWE – HC: $p = 0.016$, $CI_{95} = [-8.006_{e-12}, -1.759_{e-12}]$) similar to what we have found at occipital and parietal electrodes. This is most likely the result of an overall increase in activity in the PWE, not specific to the PPR band, as argued in the main text. All comparisons are shown in Figure 1C&D.

**6.2. Frontal PPR power in comparison to PWE**

Pairwise comparisons of PSE groups to PWE revealed, comparable to the parietal and occipital electrodes, a significantly increased frontal PPR power in PSE patients, particularly in Type 4 for stimulation frequencies 10 Hz (Type 4 – PWE: $p = 0.006$, $CI_{95} = [-3.134_{e-10}, -8.854_{e-11}]$), 15Hz (Type 3 – PWE: $p = 0.036$, $CI_{95} = [-1.884_{e-10}, -9.823_{e-12}]$; Type 4 – PWE: $p = 0.006$, $CI_{95} = [-2.379_{e-10}, -5.987_{e-11}]$), 20Hz (Type 4 – PWE: $p = 0.006$, $CI_{95} = [-2.279_{e-10}, -9.422_{e-11}]$), and 25Hz (Type 4 – PWE: $p = 0.006$, $CI_{95} = [-2.444_{e-10}, -4.684_{e-11}]$). For stimulation frequency 5 Hz, all $p > 0.971$. Similar to parietal electrodes, Type 2 and Type 3 showed no significant difference in frontal PPR power.

**6.3. Frontal PPR power in comparison to Type 4**

Finally, we compared PSE groups to Type 4. It revealed the same pattern of PPR power increase with type severity as in parietal and occipital channels. Type 4 showed significantly increased power compared to other PSE types for stimulation frequencies 10 Hz (Type 2 – Type 4: $p = 0.004$, $CI_{95} = [-3.284_{e-10}, -1.065_{e-10}]$; Type 3 – Type 4: $p = 0.036$, $CI_{95} = [-3.016_{e-10}, -5.989_{e-11}]$), 15 Hz (Type 2 – Type 4: $p = 0.004$, $CI_{95} = [-2.379_{e-10}, -5.439_{e-11}]$), 20 Hz (Type 2 – Type 4: $p = 0.004$, $CI_{95} = [-2.282_{e-10}, -1.008_{e-10}]$; Type 3 – Type 4: $p = 0.004$, $CI_{95} = [-1.975_{e-10}, -7.166_{e-11}]$), and 25 Hz (Type 2 – Type 4:

$p = 0.012$, $CI_{95} = [-2.331_{e-10}, -4.165_{e-11}]$; Type 3 – Type 4: $p = 0.036$, $CI_{95} = [-2.352_{e-10}, -4.063_{e-11}]$). For stimulation frequency 5 Hz, all $p > 0.152$. All reported p-values are Bonferroni corrected.

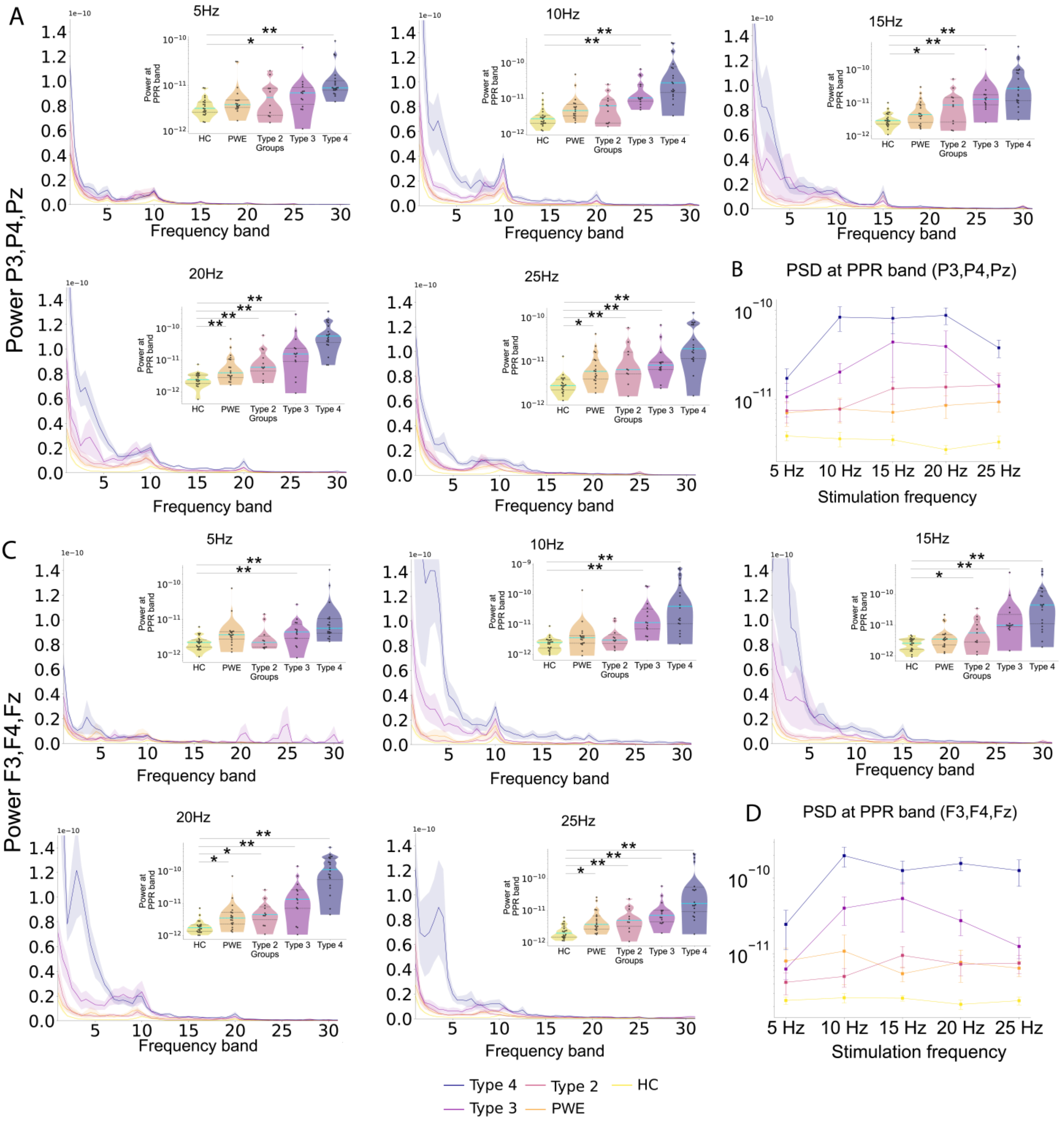


**Figure 1.: Spectral power differences at parietal and frontal electrodes compared to HC.** (**A, C**) Power spectral densities (PSDs) displayed for each stimulation frequency averaged per group (**A**) at parietal electrodes (P3, P4, Pz), (**C**), and at frontal electrodes (F3, F4, Fz). PSDs show power fluctuations at the bands linked to the triggering frequency and the pathological PPR activity between 3-4 Hz (PPR band) of PSE patients. The shaded area shows the standard error corresponding to each group's average. Stimulation frequency-specific PPRs are further displayed on the violin plots. Significant differences between HC and the patient groups are displayed. Statistical significance is represented as $*p \leq 0.05$, $**p \leq 0.01$ and $***p \leq 0.001$. (**B, D**) The line plot displays the average power, with standard error, for all. It shows an increase in PPR activity for stimulation frequencies previously shown to be high risk (10, 15, and 20 Hz) and still high but decreasing PPR for 25 Hz. Both the violin plots and the line plot show log-transformed values.

### 7. Functional connectivity in comparison to PWE

To further delineate differences in FC of PSE and non-PSE patients, PLV values of PSE patients were analyzed compared to PWE. We observed a significant increase between frontal regions and frontal and occipital regions in PSE patients compared to PWE. Increased FC between frontopolar and frontotemporal regions emerged for stimulation frequencies 10 Hz (Type 3 – PWE: p = 0.006; $CI_{95}$ =[0.007, 0.157]), 15 Hz (Type 3 – PWE: p = 0.012; $CI_{95}$ =[0.008, 0.158]; Type 4 – PWE: p = 0.018; $CI_{95}$ =[0.007, 0.159]), and 20 Hz (Type 3 – PWE: p = 0.006; $CI_{95}$ =[ -0.007, 0.163]). Anterior FC was also shown to be strong between frontopolar and frontocentral and between frontotemporal and frontocentral electrodes, illustrated in Figure 3. While anterior to posterior FC also increased in Type 3 between frontotemporal and occipital electrodes for stimulation frequency 10 Hz (p = 0.024, $CI_{95}$ = [-0.047, 0.100]), and in Type 4 compared between frontocentral to occipital for stimulation frequencies 10 Hz (p = 0.006, $CI_{95}$ = [-0.070, 0.097]), 15 Hz (p = 0.018, $CI_{95}$ =[-0.089, 0.058]), 20 Hz (p = 0.018, $CI_{95}$ = [-0.085, 0.063]). A decreased FC between central and parietal and central and frontal regions also emerged, although to a lesser extent than when the groups were compared to HC. Most of the hypoconnectivity between these areas is only visible on the topoplots showing between channel connectivity but does not appear as significant on the region-wise plot (Figure 3). Significantly decreased FC was found between central and frontopolar regions for stimulation frequencies 10 Hz (Type 3 – PWE: p = 0.006, $CI_{95}$ = [-0.029, 0.132]), and between central and frontocentral regions again for 10 Hz stimulation frequency (Type 2 – PWE: p = 0.006, $CI_{95}$ = [-0.066, 0.019]). The rest of the FC differences are illustrated in Figure 3.

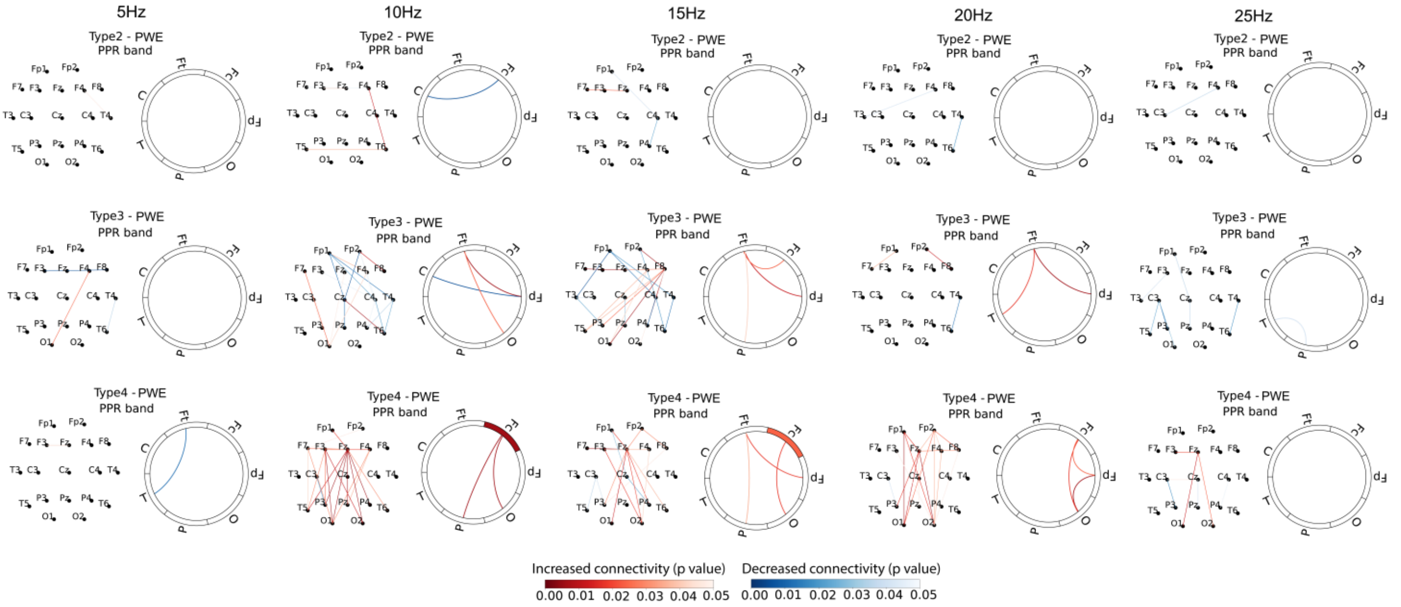


**Figure 3: PSE FC patterns in comparison to PWE.** Significant functional connectivity patterns are shown channel (topo plots) and region-wise (spider plots) for all five stimulation frequencies. The red color indicates increased, while the blue color indicates decreased connectivity in PSE patients compared to PWE. This was calculated by subtracting PWE from the respective PSE patient group (Type 2, 3, and 4). The shade of the colors corresponds to the level of significance, with darker colors indicating higher significance. In the case of a significant within-region connectivity difference, the level of significance and the nature of change (decrease, increase) are indicated by the colored in section at the corresponding region of the spider plot. Hyperconnectivity shows enhancement between occipital and frontal and within frontal regions with type severity. Hypoconnectivity is present only on the channel level between central and parietal and between central and frontopolar channels PWE in all PSE types, but does not appear on the region-wise level.
